# Learning-by-Doing to Adapt a Digital Community Engagement Strategy for Identifying and Reaching Zero-Dose Children in Kano State, Nigeria: A Program Case Study

**DOI:** 10.64898/2026.10.08.26365040

**Authors:** Amina Ahmad K U Nagogo, Nafisa Sani Nass, Amina Muhammad, Rahimat Abubakar, M.B Nurudeen, Yahaya Mohammed, Shamsudeen Sani, Jenny Sequeira, Patrick Y Liu, Katie Maloney, Abdulmumin Saad, Yusuf A Yusufari, Salisu Ahmed Ibrahim, Nancy Fullman

## Abstract

**Background:** Kano State has one of the highest concentrations of zero-dose (ZD) children in Nigeria, challenging equitable immunization coverage. Digital engagement tools meant to strengthen timely data to address such gaps often underperform due to limited adaptability to user needs. Learning-by-doing (LxD), which brings together human-centered design and adaptive management, offers more context-responsive solutions.

**Intervention and Implementation Approach:** MCH-Track, a digital platform for tracking and referral for childhood vaccinations was used by 56 female community mobilizers in two local government areas (LGAs) out of the 44 LGAs in Kano State, Nigeria. We conducted a program case study from: 1) August 2024-April 2025 (initial implementation); and 2) adapted implementation (May 2025-November 2025), where LxD was incorporated to address emergent challenges. LxD included structured shadowing, monthly interviews with users, after-action reviews, and change log to track adaptations alongside routine services. Performance was monitored using four indicators: feedback actioned, response time-to-user feedback, ease-of-use score, and mobilizer performance in tracking ZD children.

**Findings:** After incorporating LxD, 17 issues were identified and led to 12 prioritized adaptations. All logged issues received responses, with median response time declining to approximately two weeks, while community mobilizers reported higher usability of the digital platform (e.g., mean ease-of-use score of 4.1 out of 5). Enhanced root-cause analysis enabled trend tracking and clearer escalation pathways for systemic issues like micro-stockouts at immunization points. Empathy tool records revealed mobilizer struggles with platform. This was immediately logged, prompting user experience redesign into a seamless, single-step process from registration-to-referral. During the LxD implementation period, an average of 33 ZD children per month were identified via MCH-Track, a substantial increase from the pre-LxD period (an average of 11 ZD children identified per month). Average referral-to-access rates also improved during the LxD period, rising from 44% of identified and referred ZD children accessing immunization services pre-LxD to 70% after incorporating LxD practices.

**Conclusion:** Embedding LxD within MCH-Track strengthened co-creation, streamlined workflows, and improved community mobilizer performance, demonstrating the value of adaptive management in reaching ZD children.

**Teaser Message:** Integrating learning-by-doing approaches - mixture of human-centered design and adaptive management - into a digital community-engagement platform improved understanding of previously under-recognized vaccination barriers, implementing locally tailored adaptations, and strengthening frontline responsiveness to reach zero-dose children in Kano State, Nigeria.

**Key Messages:** *Key Findings:*

- Co-creation with community mobilizers and transparent change log use translated frontline feedback into trackable system improvements, improved birth-registration-to-referral workflows, and enhanced identification and follow-up of zero-dose children.
- Embedding iterative rapid cycles within the digital community-engagement platform addressed weak feedback loops and unified decisions between mobilizers and supervisors.
- Incorporating monthly learn-by-doing cycles into MCH-Track implementation strengthened how community mobilizers helped households overcome vaccination barriers through root-cause-analysis. For instance, insights from After-Action Reviews revealed “programmatic mistrust” was linked to “male-resistance”, a key barrier to vaccination.

*Key Implications:*

- Health system managers and implementing partners could benefit from incorporating learn-by-doing approaches - human-centered design plus adaptive management – into existing platforms (e.g, Health Partners Forums) and prioritizing co-creation with communities who face challenges with services, gender dynamics, and trust.
- Program managers can combine digital tools with structured feedback cycles to better identify and respond to healthcare worker needs, increase efficiency and quality.
- Embedding adaptive learning processes within routine immunization governance and financing structures could promote more responsive and resilient programming.

## Background

Despite substantial global progress in childhood immunization, zero-dose (ZD) children - often represented by infants who have infants who have not received the first dose of diphtheria, tetanus, and pertussis-containing vaccine (DTP1) by the end of their first year of life - remain concentrated in fragile, underserved, or marginalized communities.^1^ Nigeria has one of the largest ZD populations worldwide, approximately 2.1 million ZD children in 2024.^2^ As Nigeria’s most populous state, Kano has among the highest ZD prevalences in the country, with estimates reaching 45%.^2^ Located in northern Nigeria and considered patriarchal, Kano State has experienced persistent routine immunization gaps related to geographic access and supply constraints, as well as interacting social, behavioral, and health system factors. Such barriers to vaccination include inequitable decision-making power (male dominance);, misinformation about immunization, fear of side effects, service experience challenges (e.g., long wait times), and health system constraints such as vaccine stockouts.^3^ Conventional reporting and monitoring systems, such as the District Health Information System (DHIS2), mostly capture quantitative data such as vaccine doses administered and rarely reflect challenges experienced by ZD caregivers or frontline health workers. Because routine immunization (RI) managers largely rely on such quantitative data systems to inform programming, lacking more detailed and actionable insights around root causes of ZD likely affects program reach and effectiveness.^1^

Digital community-engagement platforms offer new opportunities to strengthen enumeration, referral, and follow-up of ZD children.^4^ However, digital tool implementation often involves static program designs that insufficiently respond to evolving frontline realities, usability challenges, and health system bottlenecks.^5^ Community health worker–facing digital tools show potential to improve access and client health seeking behaviors but rely heavily on sufficient training, supervision, and supportive health system structures to achieve meaningful impact. ^6^ As a result, the potential for digitization to improve equitable immunization coverage is frequently undermined by weak feedback loops and slow adaptation.^4, 5^ Emerging evidence from the WHO African Region highlights the importance of adaptive learning systems, which use continuous, feedback-driven processes to generate context-specific insights and guide real-time programmatic adjustments in complex immunization environments.^7^

Learning-by-doing (LxD) refers to structured, iterative implementation approaches that integrate principles of human-centred design (HCD), continuous quality improvement (CQI), and adaptive management to enable co-creation with end users and rapid feedback cycles (Box 1) ^8^ Through this process, LxD generates real-time insights and continuously refines both problem identification and intervention delivery in response to evolving implementation challenges^.8^ When embedded within digital community-engagement systems, this approach can enhance frontline responsiveness, strengthen community trust, and improve identification and outreach to ZD children by shortening feedback-to-action cycles and translating frontline insights into timely, context-specific programmatic adaptations. ^8^ However, empirical evidence on the application of LxD within RI digital platforms remains limited.

This program case study describes the implementation and early outcomes of an LxD-guided adaptation of a digital community-engagement intervention under the Zero-Dose Learning Agenda (ZDLA) in Kano State, Nigeria (Box 2). We focus on a subset of early outcomes following LxD incorporation, including community mobilizer ease-of-use metrics and community mobilizer performances in tracking and referral of ZD children for immunization services.

### Box 1. Learn-by-doing: where human-centered design and continuous learning connect

Learn-by-doing (LxD) is a general term for a set of approaches that can be used to better understand and address persistent health system challenges, supporting more flexible, efficient, and equitable health service delivery.

LxD embeds human-centered design, adaptive management or continuous quality improvement, and gender-responsive practices in routine health system workflows to improve processes, products or tools, and planning at different levels of the health system. End users, such as caregivers, community members, and frontline providers, work together with health program implementers to:

- Identify actionable drivers and root causes;
- Co-design interventions to ensure feasibility, relevance, and ownership; and
- Continuously review and adapt interventions to identify what is working, what is not, and how they can be improved.

### Box 2. Zero-Dose Learning Agenda: a 2-year initiative to better understand how to reach and reduce zero-dose children

The Zero-Dose Learning Agenda (ZDLA) was a 2-year learning initiative by the Gates Foundation to better understand how to accelerate progress toward the IA2030 goal of achieving a 50% reduction in ZD. Through ZDLA, partners applied learn-by-doing (LxD) techniques and practices in high-ZD settings to:

- Diagnose drivers of ZD and understand their root causes;
- Work collaboratively with end-users, including caregivers, community leaders, healthcare workers, and government officials, to co-design, implement, and iteratively adapt interventions;
- Reflect gender-related barriers and responsiveness; and,
- Capture costing information.

This two-year initiative (2023–2025) operated across select subnational areas in DRC, Ethiopia, India, Kenya, Nigeria, and Pakistan.

### Intervention and implementation approach

#### Intervention overview

From August 2024 through November 2025, we, Datharm’s program staff implemented an intervention known as Maternal and Child Health Track (MCH-Track),^9^ a mobile-enabled digital community engagement platform designed to strengthen household-level enumeration, referral, and follow-up of ZD children.^9^ Box 3 summarizes the MCH-Track intervention as originally developed by Datharm and the Kano State Primary Health Care Management Board (KSPHCMB), as well as its defaulter tracking application.

##### Box 3. MCH-Track intervention: original design and features

The MCH-Track digital platform (Figure 1) was launched in 2024 to streamline community engagement strategies, increase efficiency and address lingering issues such as time-intensive reconciliation of data, fragmented referral tracking, and increase visibility of ZD children and others lacking access to timely maternal and child health services. poor visibility of defaulters.

**Figure 1.**
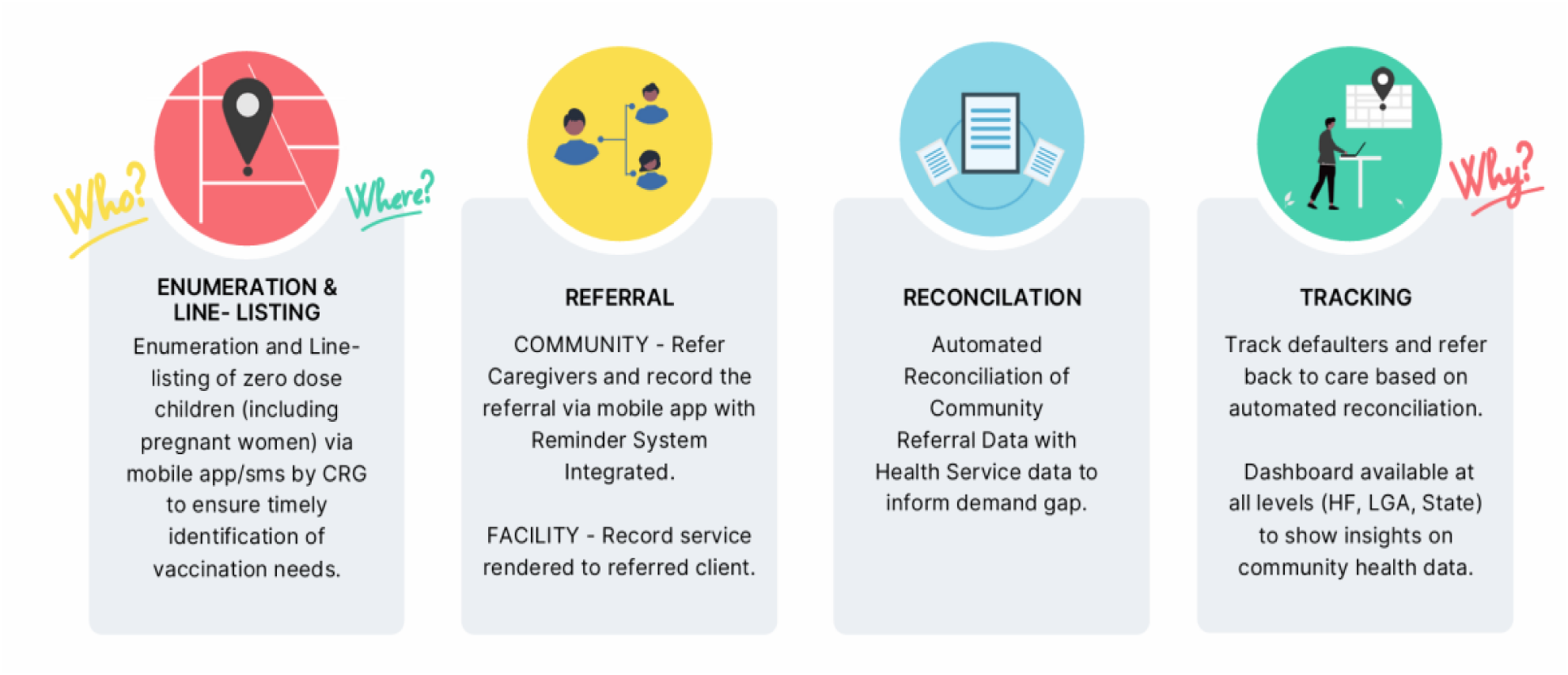
High-level pillars of MCH-Track.

MCH-Track is grounded in the following theory of change (Figure 2): systematically capturing real-time, community-generated data and using those data to provide linkages to service delivery can improve the identification of ZD children, strengthen coordination between community actors and primary healthcare (PHC) facilities, and support timely follow-up and service utilization.

**Figure 2.**
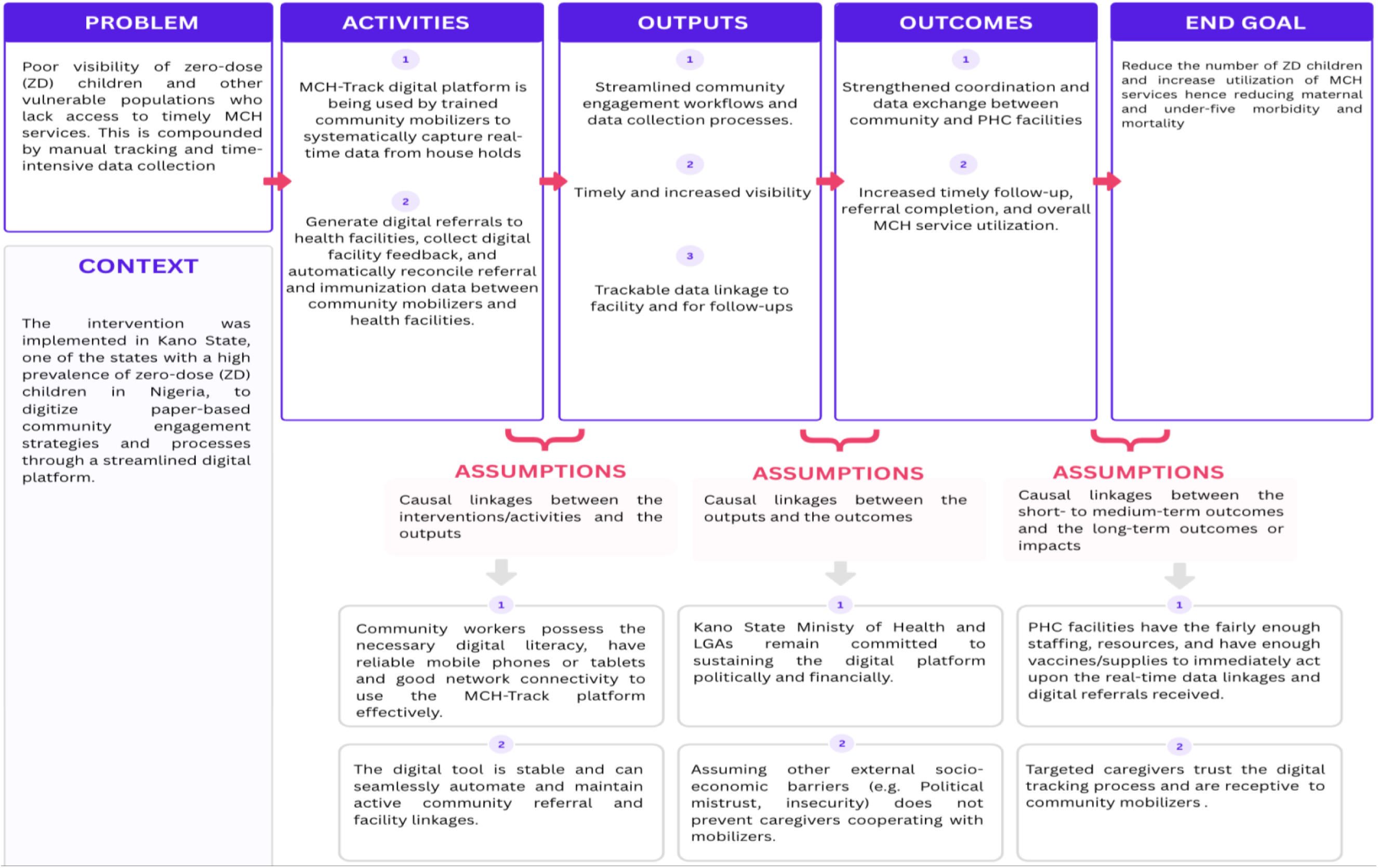
Theory of Change for MCH-Track implementation.

MCH-Track was originally designed to support a closed-loop referral system (Figure 3), as illustrated by steps 1-7 in the illustration below:

- Step 1: Community mobilizers use the digital platform to conduct household-level enumeration and line-listing, including registration of pregnant women and births as well as identification of and referrals for children eligible for immunization;
- Step 2: PHC facilities document services provided;
- Steps 3, 4, and 5: An automated reconciliation process links community referrals with facility service data to identify missed opportunities and defaulters;
- Steps 6 and 7: Interactive dashboards at facility, local government area (LGA), and state levels support onward decision-making and action by immunization managers.

**Figure 3.**
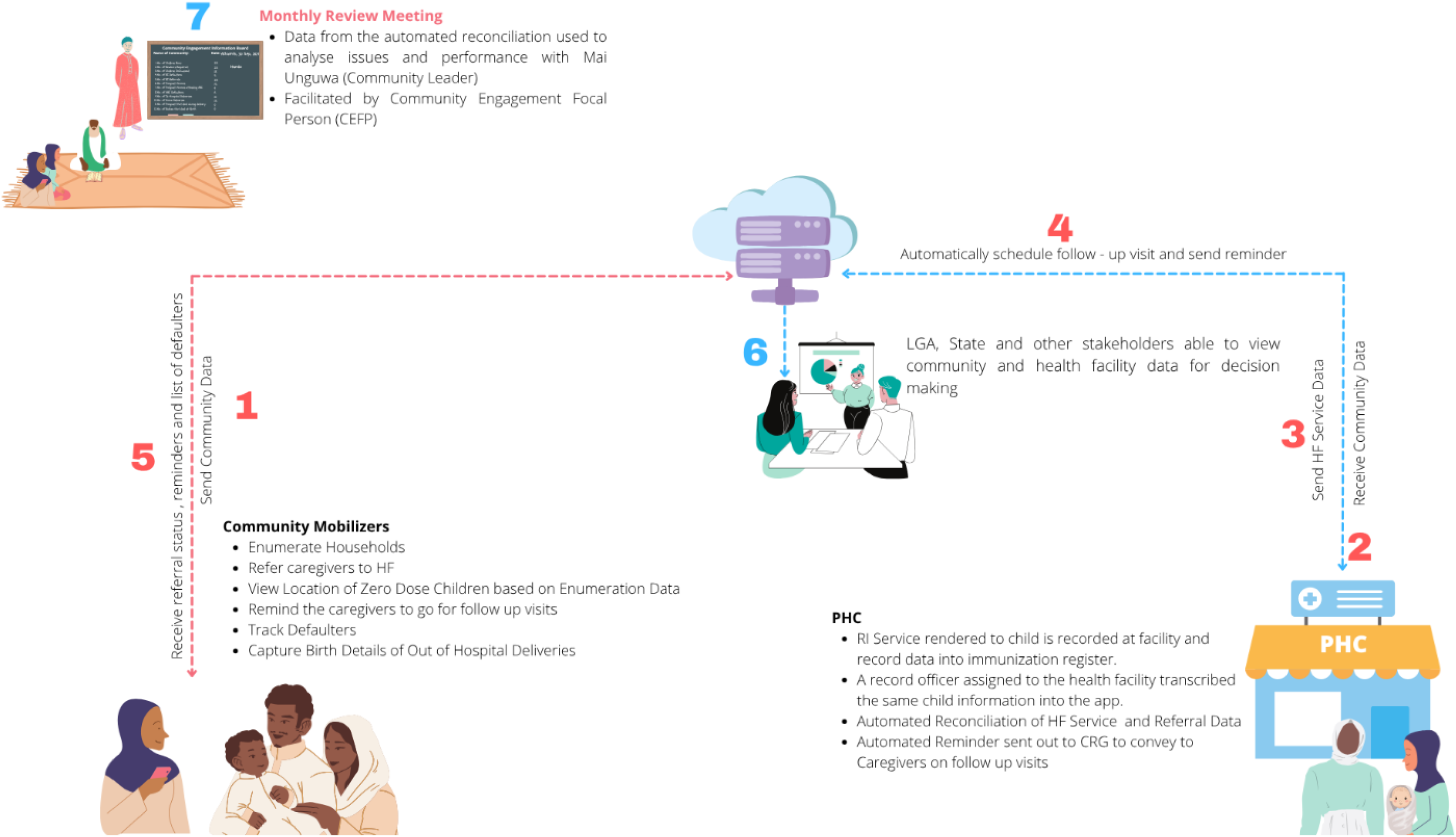
Visual depiction of MCH-Track’s closed-loop referral system.

MCH-Track also allows community mobilizers to capture caregiver-reported reasons for non-vaccination through its defaulter tracking application. The original design included 13 response options based on access and intent consisting of no felt need for vaccination, had family problems (personal illness, death in family etc), lack of adequate means of transportation, lack of adequate financial resources, missed scheduled vaccination days, information gaps/lack awareness of vaccination schedules, caregiver being busy, fear of adverse events following immunization (AEFIs), obtaining permission from parent or guardian, healthcare provider is not of the same sex, poor attitude of health providers, uncooperative husbands, low trust in vaccines.^10^

A total of 56 community mobilizers (active in study sites) from the cohort of 274 female community volunteers in the two LGAs were involved in the implementation of MCH-Track. These mobilizers are serving as co-creators of the intervention and operating as the connectors between primary healthcare (PHC) facilities and communities to support the identification, registration, referral, and follow-up of children at risk of missing vaccination. In this context, community mobilizers are similar to indigenous community volunteers; they are locally embedded, female-led groups focused on community engagement and mobilization for primary health care services. Only 48% of which were engaged by the State Government to support local immunization strategies, operating on a monthly stipend of USD 3.80.

MCH-Track was implemented in 20 communities (10 each) in Ungogo and Gabasawa Local Government Areas (LGAs) of Kano State, Nigeria selected purposively from 44 LGAs of Kano State. These LGAs include both urban (Ungogo) and rural (Gabasawa) communities that are characterized by persistent inequities in routine immunization coverage and a high burden of ZD children (Table 1).^11^

**Table 1.** Summary characteristics of MCH-Track implementation areas and intervention indicators. Estimated catchment populations and households or children reached by the intervention were based on MCH-Track line list enrolment figures from 2025.

| Indicators |  | Ungogo LGA | Gabasawa LGA |
| --- | --- | --- | --- |
| Select site characteristics | Setting type | Urban;<br>high-population density | Rural;<br>scattered populations |
|  | Estimated DTP1 coverage (%) [12] | 69% | 57% |
|  | Estimated ZD prevalence (%) [12] | 31% | 43% |
| Select MCH-Track intervention indicators | Community mobilizers | 29 | 27 |
|  | Health facilities | 11 | 10 |
|  | Estimated catchment population, households | 19,459 | 8,468 |
|  | Estimated catchment population, individuals enrolled | 21,494 | 12,651 |
|  | Estimated households reached with children aged < 2 years | 4,194 | 5,394 |
|  | Estimated children aged < 2 years reached | 8,004 | 7,137 |

### Intervention adaptation and learn-by-doing implementation

After nine months of intervention implementation (August 2024-April 2025), we conducted a needs assessment. Several process indicators showed positive results for this initial implementation period, including the identification and line-listing of 7,210 children and 2,187 women. At the same time, several implementation challenges emerged. First, despite having access to the platform’s interface, some mobilizers struggled to use it when collecting data from caregivers in the filed – a signal of potential ease-of-use problems and/or workflow burdens. Second, feedback loops between community mobilizers and software developers experienced frictions, with community mobilizers regularly raising challenges and developers only addressing them on a case-by-case basis. Responses to deeper issues (e.g., workflow problems) were often delayed, leading to frustration for community mobilizers. Lastly, frontline ownership of program implementation was limited, especially since community mobilizers experienced minimal agency around how the platform worked.

To address these implementations challenges, we sought to redesign key aspects of the intervention and its processes using LxD approaches, of which were also being implemented elsewhere in Kano and globally.^13^ At a high-level, we applied an iterative, four-stage LxD cycle: **1) Reflection and documentation** of frontline insights**; 2) Review**, whereby emerging root causes and patterns are collaboratively reviewed and prioritized; **3) Reaction,** such that locally-tailored adaptations are implemented; and **4) Redeployment**, which involved redeploying and reassessing updated workflows in subsequent cycles. From May 2025 to November 2025, we conducted four of these learning cycles, of which were informed by several LxD practices and tools; we detail them in Box 4.

#### Box 4. Learn-by-doing (LxD) practices and tools implemented for MCH-Track

##### Co-creation as continuous implementation practice

We embedded co-creation as a continuous implementation practice, anchored by two main user groups: 1) community mobilizers; and 2) an advisory group comprised of facility in-charges, LGA RI officers, ward focal persons, and traditional leaders. This structure ensured that adaptations were grounded in frontline realities and connected to existing RI governance systems. A participatory kick-off session involved both groups, during which they used ‘5 Whys’ to conduct root cause analyses and better understand underlying issues with the current traditional CE strategy, why ZD children were being missed, and build shared ownership of the digital intervention. To sustain co-creation, a safe space was established with dedicated learning facilitators in both LGAs forming a new norm of joint problem-solving approach between community mobilizers and developers effectively lowering hierarchical barriers.

##### Application of HCD techniques and methodologies

We used HCD approaches (e.g., iterative prototyping, site observations, qualitative interviews *via* key informant interviews [KIIs] and focus group discussions [FGDs]) with community mobilizers and healthcare workers to better understand workflow bottlenecks and previously under-recognized user needs. For example, structured shadowing and empathy mapping exercises showed that community mobilizers often managed fragmented birth-registration-to-referral processes by deploying paper-based workarounds. However, such short-term solutions hindered automated reconciliation processes and could affect follow-ups for newly registered births. This initial observation was captured in a structured empathy tool, prompting a redesign process of iterative prototyping to create a seamless single-step process for instant referrals. This co-creation process with end-users and developers not only promoted greater ownership, but also improved birth registration workflows for community mobilizers.

##### Regular participatory review and iterations

Third, we supported more regular touchpoints for participatory review and subsequent iterations. Members of the implementation team began conducting weekly supervisory field visits, targeting specific wards within each LGA on a rotating basis. Learning was further supported and sustained through monthly interviews, group discussions, structured shadowing, redeployment reviews, and after-action reviews (AARs) embedded within LGA routine Technical Working Group (TWG) meetings. These review meetings were held monthly and brought together community mobilizers, data officers, LGA teams, and members of the implementation team to collectively review findings, discuss issues, and agree on adaptations.

### Data sources and data collection procedures

#### Pre-LxD

Prior to integrating LxD, data collection was structured as a conventional, real-time monitoring process. Quantitative data was primarily sourced from the MCH-Track digital platform, this platform continuously streams backend analytics on indicators such as ZD children identified and referred. Data was also from facility immunization registers, while qualitative data was unstructured and reported via phone calls, Whatsapp messages or during supervisory visits on the field. Documentation of issues or resolutions by developers and supervisors was not systematic and often in phone records and written notes with some actions remaining undocumented.

#### During LxD

Quantitative data originated from the MCH-Track digital platform records and from administration of a short electronic survey questionnaire to community mobilizers to determine platform usability scores based on a 5-point Likert scale. Data was as well from existing routine immunization (RI) monitoring data (i.e., health facility immunization registers). Pre-defined implementation indicators on feedback responsiveness and issue resolution time were collected from the implementing team’s administrative records and qualitative records.

Qualitative data collected largely drew from the LxD process, which was structured as an iterative, continuous loop that generated feedback for learning and improvement. Initially, a root cause analysis (RCA) using the “5 whys” was applied on 30^th^ May 2025 during the LxD stakeholder workshop with 18 people (6 community mobilizers, 4 RI incharges,4 community leaders, one LGA RI manager and one Supportive Supervision Program Officer to explore the root causes of persistent zero-dose children and why facility community review meetings were not being held.

##### Key Informant Interviews (KIIs)

Four KIIs were conducted, two with community mobilizers from each LGA and two with RI in-charges from Gabasawa LGA mainly to clarify bottlenecks with MCH-Track usage.

##### Focus Group Discussions (FGDs)

FGDs became formal reflection sessions. Nine FGDs were conducted in Gabasawa and two in Ungogo. The number of sessions was based on data saturation, as rural Gabasawa presented more issues for probing. An FGD guide translated in Hausa language was used, and each session was recorded and noted with groups comprising of up to 10 community mobilizers.

##### Day-in-Life-Shadowing

During regular supervisory visits with or without the State team, at least 3 Datharm’s staff members observed all 56 community mobilizers in both LGAs periodically and noted user experiences. This uncovered discrepancies what challenges or gaps exist within the platform that would need fixing. These shadowing exercises were conducted at least twice a month. Empathy Mapping based on data from the Shadowing activities conducted was used to summarize emotional and behavioral insights from the users’ point of view categorized into “Says, Feels, Does, Thinks”.

Above processes provided data within the reflection and documentation phase. Data gathered from reflections were used for review in the Learning Logs which serve as structured documents for spontaneous field insights, unexpected technical glitches and user suggested fixes or ideas. Whenever a learning log helped identify a flaw, the modifications to be made to the MCH-Track or an activity, due dates and status of change were recorded on a Change Log to preserve traceability.

We operationalized continuous adaptation through two “living documents”: 1) a Learning Log; and 2) a Change Log **(Table 2).** In brief, the Learning Log served as the institutional memory across cycles, reflecting feedback collected across users, ideation around potential solutions to challenges and their feasibility, and subsequent decisions or actions to take. The Learning Log was regularly reviewed by the implementation team and was a primary input for determining which issues were escalated to the Change Log. The latter served as an internal record to formally track prioritized issues through their resolution **(Figure 4).**

**Figure 4.**
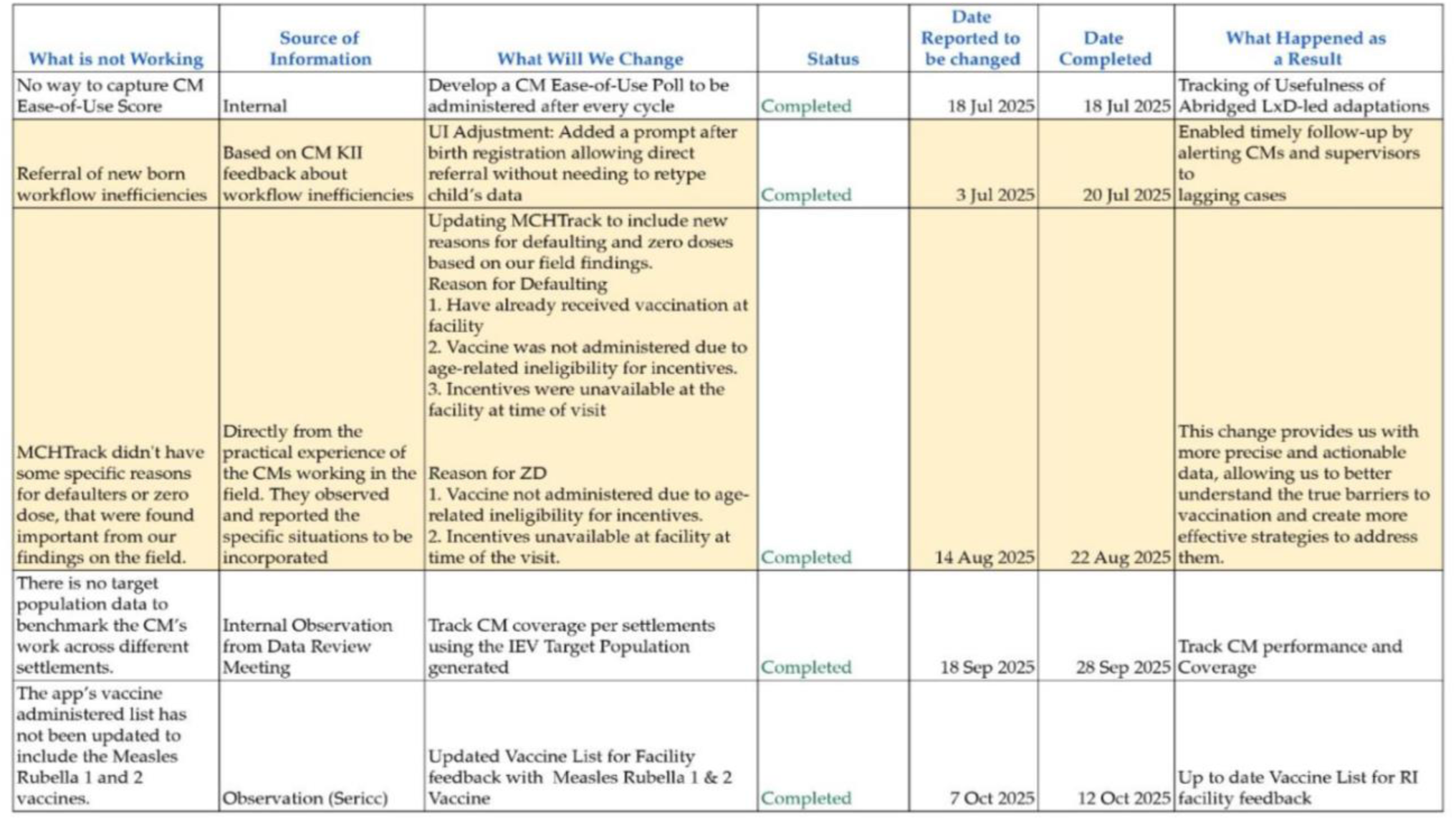
Snapshot of sample excerpt from the MCH-Track Change Log.

**Table 2.** Learning Log and Change Log applications for MCH-Track.

|  | **Learning Log** | **Change Log** |
| --- | --- | --- |
| **Summary of main use(s)** | • Primary means of documenting key successes, needs, and challenges around intervention implementation over time<br>• Key input for review meetings and internal planning sessions to help with prioritizing issues for escalation into the change log | • Primary means of documenting adaptations, actions taken, implementation status, and accountability for translating feedback from the Learning Log to concrete improvements |
| **Summary of data collected** | • Frontline observations about contextual triggers and user sentiment (e.g., frustration, discouragement, motivation) during implementation<br>• User-reported challenges and underlying causes<br>• User-reported suggestions for potential fixes or solutions (and feasibility ratings)<br>• Decisions made | • Prioritized issue for resolution<br>• Action plan for resolution<br>• Target date of resolution<br>• Outcomes |
| **User groups represented** | • Community mobilizers<br>• Data officers<br>• Facility staff | *None*; internal implementation team tool for accountability and progress tracking |
| **How the data were collected** | • Direct feedback from user groups<br>• Supervisory visits at facilities<br>• Interviews and FGDs<br>• Review meetings | • Inputs from the Learning Log<br>• Implementation team updates<br>• Observation |

### Data analysis

Qualitative data were synthesized using implementation-focused thematic analysis. Interview and discussion data were transcribed, reviewed, and coded using a combination of inductive and framework-informed approaches aligned with implementation objectives. Codes were iteratively refined and grouped into themes reflecting key implementation barriers, facilitators, and adaptation processes. For quantitative data, we generated descriptive statistics to assess implementation outcomes, based on the following indicators: ease-of-use scores, feature/adaptation usefulness ratings, percentage of feedback actioned, median response time to user feedback, and measures of community mobilizer motivation and ownership.

### Ethical Considerations

Ethical approval was obtained from the Kano State Ministry of Health Research Ethics Committee (NHREC/17/03/2018) as part of a larger study. All participants provided informed consent. Qualitative and programmatic data were de-identified, and data-protection safeguards, including removal of personal identifiers, role-based access to digital data collection platforms, password-protected databases, and secure storage on encrypted servers, were maintained throughout implementation and analysis.

### Implementation findings

#### Intervention implementation adaptations with learning-by-doing

From May 2025 through November 2025, we identified 17 distinct field implementation issues, with 12 prioritized for adaptation and 11 with completed resolution at the time of reporting. At a high level, identified issues largely fell within two main categories: challenges directly related to intervention implementation and challenges more related to implementation context or broader community engagement:

##### Intervention implementation challenges

1) A disjointed digital workflow that required users to complete newborn line-listing/birth registration and referral for birth-dose immunization as separate steps, increasing time and data entry burden.; and
2) Gaps in understanding reasons for non-vaccination, due to limited probing options and fields on the digital tracking tool (e.g., stockout of other vaccination commodities other than vaccines and “no reason” as an option which could mean concealment of reasons by caregivers).

##### Implementation context and community engagement challenges

3) **Low community trust in the vaccination program,** with ZD household experiences of feeling excluded from government benefits emerging as an important root cause for non-vaccination;
4) **Traditional leaders not included in monthly LGA TWG meetings**, where AAR-based findings, priorities, and adaptations were discussed; and
5) **Limited number of community mobilizers relative to catchment area populations,** with minimal support and incentive structures.

Based on user feedback and prioritization, we implemented three main adaptations **(Table 3).** These intervention adaptations, as well as broader implementation of LxD processes, were associated with higher platform usability, feedback responsiveness, and frontline ownership of routine immunization activities **(Table 4).** While there were frequent reports of frictions prior to implementation, the mean ease-of-use score of the MCH-Track improved to a score 4.1 out of 5 among community mobilizers, and average response time from problem identification to resolution decreased from about 4 weeks to ≤ 2 weeks. The proportion of ZD children referred to those identified decreased slightly by percentage points. This decrease may be related to increases in the total number of ZD children referred after LxD processes were implemented, while the number of community mobilizers and other resources remained constant. There was a 44% referral-to-access rate pre-LxD and 70% rate during the LxD period (26 percentage point difference). More defaulters were also tracked and referred for immunization within the LxD period, increasing from an average of 149 per month pre-LxD to an average of 281 per month during LxD implementation (a 89% rise) **(Table 4).**

**Table 3.** Three MCH-Track intervention adaptations.

| **Adaptation area** | **Referral workflow** | **Defaulter tracking** | **Community Engagement** |
| --- | --- | --- | --- |
| **Identified problem** | High workflow burdens experienced by community mobilizers | Limited understanding of reasons for non-vaccination | Low community engagement and programmatic trust |
| **Underlying cause** | Fragmented birth-to-referral processes in the module, resulting in HCWs having to exit the application between entering birth registrations and initiating referrals<br>“If it is possible like when it comes to the birth registration, it should be easy to refer a child after doing their birth registration because presently, you would have to go back and look for their parents' names. If it is possible, we should be able to link the referrals to where we did the child's birth registration and just send it, that way you do not have to go back and start a new process.” – Community mobilizer, Ungogo | Defaulter tracker included a limited number of predefined reasons for non-vaccination (e.g., missed appointment, lack of awareness), and did not reflect a wider range of challenges or needs reported by community mobilizers (e.g., incentives not being available, caregivers being ineligible for incentives, service disruptions) | Limited inclusion of traditional leaders in typical planning processes and low (or no) regular community-facility feedback mechanisms to better respond to local and/or emergent challenges around vaccination (e.g., challenges associated with incentives, supply gaps, and gendered constraints around decision-making and agency) |
| **Implemented change** | Added automatic referral prompts immediately after registration, eliminating process gaps | Expanded defaulter-reason fields based on community mobilization feedback and after-action reviews. This includes reasons such as “no incentives”<br>Included referral outcomes within defaulter application | Supported structured dialogues with facility in-charges and traditional leaders, supported by dashboard insights and after-action reviews |
| **Outcomes** | Streamlined workflow, reduced backtracking and re-entry, reduced time burdens, improved data completeness, and higher user confidence | More actionable insights around caregiver-experienced barriers to vaccination and subsequent linkages to issue resolution<br>*Example:* Reclassified “missed appointment” to more distinct categories such as “incentive unavailable” and “ineligible for incentive”, which then supported more targeted responses to system and/or community needs | More locally-tailored engagement, strengthened accountability for service delivery, and building greater trust with community leaders |

**Table 4.** Implementation outcomes before and after implementing LxD adaptations.

| **Implementation indicators** | **Initial implementation (Pre-LxD)**<br>**(August 2024-April 2025)** | **Implementation with LxD approaches and adaptations (May 2025-November 2025)** |
| --- | --- | --- |
| **Platform usability** | Reported workflow burden and friction | A mean usability score of 4.1 out of 5 among community mobilizers<br>(*n*=42, using a Likert scale of 1 being very difficult to use and 5 very easy to use) |
| **Feedback resolution** | Responses to frontline feedback were often delayed, undocumented, or inconsistently acted upon, with no formal mechanism to track whether reported issues resulted in programmatic changes | All (100%) of reported issues addressed |
| **Response time** | Prolonged resolution cycles of $\geq 4$ weeks | $\leq 2$-week median resolution time |
| **Community mobilizer ownership** | Frustration and low ownership<br>“...you would have to go back and look for their parents’ names... and start again.”<br>(Community Mobilizer, Rural)<br>“If it is possible... we should be able to link the referrals... so you do not have to go back and start a new process.” (Community Mobilizer, Urban) | Substantial increase in confidence and ownership because of the bottom-up approach during the process of LxD. The change log especially reinforced the principle that feedback leads to action, fostering a culture of agency and shared ownership. |
| **Total ZD children identified (average per month)** | 98 (10.9 per month) | 232 (33.1 per month) |
| **Total identified ZD children with at least one referral (average per month)** | 82 (9.1 per month) | 186 (26.6 per month) |
| **Total referral rate (% of identified ZD children receiving at least one referral)** | 84% | 80% |
| **Total referred ZD children who accessed immunization services (average per month)** | 36 (4 per month) | 131 (18.7 per month) |
| **ZD resolution rate (% identified ZD children with referrals who accessed immunization services)** | 43.9% | 70.4% |
| **Total children (0-23 months inclusive of ZD) referred to immunization services (average per month)** | 5499 (611 per month) | 8354 (1193 per month) |
| **Total defaulters (0-23 months inclusive of ZD) tracked (average per month)** | 3065 (341 per month) | 7254 (1036 per month) |
| **% of defaulters (0-23 months inclusive of ZD) tracked, referred back to immunization services and vaccinated** | 25% | 29.7% |

#### Improved understanding of ZD drivers and gender-related barriers

Through intervention implementation and LxD processes like shadowing and AARs, our understanding of key drivers and root causes of ZD became more actionable and grounded in local realities. For instance, our analyses found that identified ZD children lived within an average of 1.2 km from the nearest health facility, below Nigeria’s benchmark of 2 km for fixed site service access.^14^ Accordingly, we shifted functions for MCH-Track beyond its original focus (i.e., largely referrals and overcoming access challenges) toward a system that could support more proactive identification and response to a broader range of vaccination barriers beyond access alone. We also moved from more static, high-level categorizations of non-vaccination to those more directly reflecting root causes **(Table 5),** strengthening the timely identification of key needs and corresponding responses for households with ZD or under-immunized children. For instance, in August 2025, we noticed an increase in "uncooperative husband" responses, which were directly linked to the exclusion of those communities from a concurrent Long-Lasting Insecticidal Net (LLIN) distribution. This reframed our understanding of what was driving non-vaccination, from general male gender-skewed resistance to a deeper underlying cause: programmatic distrust due to inequitable provision of health services and commodities. Second, we learned that ‘stockouts’ did not fully reflect facility supply gaps, as ‘micro-stockouts’ of specific items like BCG syringes often were the main culprits. Such "last-yard" failures posed reputational costs to the health system, eroding trust built by community mobilizers. Lastly, we gained insights into an unintended consequence of well-intentioned demand-side financing initiative (an immunization program that utilizes cash transfers to increase caregiver’s demand for immunization); community mobilizers reported caregivers refusing vaccination after they learned that their child was deemed ineligible for associated financial incentives (e.g., children 12-23 months for Penta1).

**Table 5.** Key drivers of ZD identified through intervention implementation and LxD processes.

| **Driver themes** | **Specific barriers reported** | **Example quotes from the field** |
| --- | --- | --- |
| **Programmatic mistrust** | Caregivers show resistance or negative reactions to house-to-house mobilization efforts | “The major challenge we have encountered is resistance during our house-to-house outreach efforts... even from those who were previously receptive.” (Community mobilizer, FGD - Rural) |
|  | Fathers viewed LLIN distribution campaigns as inequitable and exclusionary | “Community Mobilizers were threatened or rejected by fathers... vaccination perceived as selective pressure without benefit.” (Community Mobilizers, After-Action Review, Rural) |
| **Unintended consequences of incentives** | Caregivers did not want to seek vaccination again after not receiving expected incentives, either due to non-availability at the time of service or being told their children are not eligible | “Referred child showed up but no incentive available on that day due to officer’s absence.” (Community Mobilizers, KII feedback integrated into digital tool, Rural) |
|  |  | “Caregivers rejected vaccines when child was deemed ineligible for incentives due to age (12-23 months).” (Community Mobilizers, KII feedback integrated into digital tool, Rural) |
| **Gendered constraints on agency** | Mothers have limited agency in health-seeking decisions, control over resources for transport, and needs around their husband’s permission or accompaniment | “Some of the mothers tell us that their husbands did not allow them to receive such services... they would say they needed to inform the husbands first.” (Community mobilizer, KII, Rural) |
| **Concerns about post-vaccination side effects** | Caregivers are concerned after their children experience side-effects after vaccination, particularly as they lack management support | “One of the challenges I faced was when I try to counsel caregivers on the benefit of vaccination, they say they had previous experience with side effects like leg swelling so they don’t want to do it to want to do it again for their younger children” (Community mobilizer, KII, Rural) |
| **Health system or Service readiness gaps** | Stockouts around specific supplies (e.g., BCG syringes) and service inconsistencies erode caregiver trust in healthcare workers and broader health system | “Caregivers often refused to return to facilities due to lack of vaccines or syringes.” (Community Mobilizers, KII feedback, Rural) |

We also embedded gender-sensitive learning into ongoing implementation, to better understand the drivers of ZD and improve planning to reach ZD children before exclusion became more entrenched. Data from community mobilizers showed that ZD prevalence did not considerably differ by gender (i.e., 49% of ZD children were boys and 51% were girls), whereas gendered decision-making and constraints associated with gender norms emerged as major factors. For instance, women caregivers bear responsibility for bringing children for vaccination services, but they often lack authority, time, and resources for transportation to do so. Importantly, the AARs identified programmatic mistrust as an important underlying driver of male gatekeeping. This implementation insight prompted an adaptive response in which health facility in-charges engaged traditional leaders to facilitate community dialogue, address misconceptions about vaccination and strengthen trust in immunization services.

#### Overall intervention implementation outcomes through November 2025

Intervention implementation indicators suggest improved operational reach, responsiveness, and coordination across intervention sites. Between August 2024 and November 2025, MCH-Track recorded 15,553 children and 6,108 women line-listed, 27,083 referrals, and 17,684 health facility visits; additionally, 330 were identified as ZD out of which 167 were successfully referred to immunization services vaccinated. Qualitative findings also indicate improved identification and follow-up of ZD children, greater accountability through documented adaptations and review structures, and stronger coordination between community actors, facility teams, and traditional leaders.

##### Interest in scale-up and institutionalization

Regular reviews of digital dashboard trends have been incorporated into monthly Immunization Review Meetings in both LGAs, reflecting early signals of further LxD adoption. Furthermore, the KSPHCMB has expressed interest in adopting the digital dashboards for LGA-level monitoring beyond the two LGAs where MCH-Track was implemented. This demonstrates great commitment and shows that embedding LxD into the health system is possible. However, transitioning the current community engagement strategy for promoting PHC services including immunization from a partner-supported program environment into routine, non-project delivery poses significant systemic challenges in resource-constrained settings like Kano State. It is expected that the government would train or provide facilitators, of whom would then dedicate ample time for structured reflections and the harmonization of data into action; furthermore, it would be expected that regular operational funds for rapid adaptations would be allocated by the state government. Grafting an LxD culture into the government immunization program would mean the continuation and expansion of the program in the 2 study LGAs along with a gradual shift into the existing structures of the other 42 LGAs. This would also have to be accompanied by development of budget line with subsequent release of funds.

### Reflections from learn-by-doing implementation

While the comprehensive baseline assessment provided essential foundational insights, its pace was misaligned with the need for a HCD and rapid adaptation. Continuous learning was important and to transform learning from an abstract concept into a visible, transparent, and actionable system, empirical data must be translated into tailored communication channels that function as a "learning-by-doing" reflection loop for specific system actors. Furthermore, digital tools like MCH-Track serve as data platforms for strengthening insights into key health indicators and better understanding why clients are not seeking or accessing services. Our implementation experience shows that incorporating LxD techniques and practices into existing interventions is not only possible, but also can strengthen intervention responsiveness to community needs and support early signals of effectiveness. Key factors that contributed to successful LxD applications included the use of dedicated learning facilitators in each LGA; conducting multiple learning cycles to address end-users’ needs; and integrating AARs into existing review meeting structures at the LGA level. Box 5 highlights the key lessons learned from integrating LxD with the digital community engagement strategy.

#### Box 5. Lessons learned from incorporating learn-by-doing (LxD)

- **Prioritize agile learning tools**: The most impactful learning emerged from lightweight, embedded tools, particularly the Empathy Tool and living Learning Log.
- **Include end-users in LxD processes from the start:** Including end-users from design of the learning experience (LxD) process ensures that the final result addresses context specific problems and eliminates biased assumptions.
- **Deploy different LxD tools and techniques for different use-cases – and ensure sufficient skills and capacities in their application:** The Change Log proved most valuable for transparency and accountability, while the Empathy Tool was critical for uncovering latent needs. Monthly AARs were essential for collective sense-making but required skilled facilitation to remain effective.

**Focus on strengthening local learning cultures:** At the site level, AARs introduced a new norm of joint problem-solving between community mobilizers and health staff, reducing hierarchical barriers. The Change Log reinforced the principle that feedback leads to action, fostering a culture of agency and shared ownership.

### Study Implications and Considerations for Scale-up and institutionalization

#### Implications for Immunization Equity, IA2030, and Nigeria’s ZD Priorities

The findings help operationalize the equity ambitions of IA2030 by showing how feedback loops, root-cause analysis, and locally tailored responses can be embedded within routine immunization delivery rather than treated as abstract system ideals.^15^ In line with the IA2030 emphasis on identifying missed communities and responding to context-specific barriers, our study demonstrates how structured LxD processes (i.e., deliberately combining HCD and adaptive management) within a digital community-engagement platform can support continuous identification, follow-up, and adaptive response for underserved children. Furthermore, adaptive learning systems offer a mechanism for translating equity principles into frontline practice by continuously identifying underserved populations, understanding context-specific barriers, and tailoring programmatic responses. This is also strongly aligned with Nigeria’s national ZD programming, which prioritizes effective approaches for identifying and monitoring ZD and under-immunized children, understanding who and where they are and why they are missed, and addressing health-system barriers to their identification and measurement.^16^ Our intervention also offers a practical pathway for translating immunization equity goals into frontline practice in Nigeria by making social, behavioural, and service-readiness barriers more visible and actionable. More broadly, the findings support a shift from viewing digital health primarily as a data-capture tool toward recognizing its role as an adaptive tool within the health system. Coverage metrics remain essential, but implementation responsiveness, feedback visibility, and community trust are equally critical determinants of equitable immunization systems and may also inform responses to other complex public health challenges that require continuous learning and locally tailored action. It is also important for immunization donors to understand the complex interactions of parallel partner programs. Closer cooperation allows donors to align programs to avoid the unintended negative consequences of uncoordinated demand-side financing.

#### Implications Beyond Immunization and Broader Health System Application

While our current work focused on immunization and specifically ZD children, LxD practices offer value-add to any health area. LxD promotes direct engagement with end users to design or adapt interventions to local realities and needs of specific populations, provides flexibility to address evolving local needs as they emerge, and identifies root causes that can be used to address persistent complex health system and service experience challenges. This approach may be particularly valuable in other priority areas such as maternal and newborn health, where care-seeking behaviours, service experience, and trust dynamics strongly influence outcomes. For example, LxD could support adaptive redesign of antenatal care pathways, improve referral linkages for obstetric emergencies, or strengthen community-facility interactions in postnatal care. Similarly, in areas such as infection prevention and control, antimicrobial stewardship, outbreak response, and primary health care service delivery, LxD offers a structured mechanism to identify root causes of persistent system bottlenecks, translate frontline insights into actionable changes, and continuously refine interventions. More broadly, these findings suggest that embedding iterative, participatory learning systems within routine health programs may represent a critical enabler for improving responsiveness, equity, and sustainability in complex, resource-constrained settings.

#### Considerations for scale-up, longer-term sustainability, and investing in LxD

For successful scale-up and sustainability, funders at subnational, national and global levels should treat MCH-Track plus LxD as an integrated operating model. This approach not only digitally identifies ZD children, but also leverages frontline feedback, root-cause analysis, and rapid adaptation to resolve reasons children remain unvaccinated. Going forward, more digital platforms should go beyond a more limited foci and take a more dynamic approach to diagnosing and responding to root causes of non-vaccination (e.g., including programmatic mistrust, micro-stockouts, unintended consequences of incentives, service experience, and gendered decision-making constraints.

Governments should also focus on making trust and service readiness measurable outcomes by embedding other quantitative key performance indicators (KPIs) into platforms such as the DHIS 2 and not just simple coverage rates. Qualitative variable measures in tools such as a “system readiness scorecard” can also be used to measure indicators such as stockout-related missed opportunities. Tracking indicators such as feedback response time, issue-resolution rates, stockout-related missed opportunities, and whether community feedback results in visible action is vital. This can be done by embedding AARs, Change Logs, Empathy Tools, and dashboard reviews into existing RI-TWG, LGA review meetings, supportive supervision, and community engagement structures. RCAs and HCD approaches can also be incorporated into annual microplanning templates to ensure interventions address local root causes and involve various groups of end-users such as traditional leaders, facility staff, and community mobilizers.

Scale-up should avoid creating donor-driven, standalone community engagement projects. Instead, LxD and digital dashboard review should be embedded within State and LGA accountability systems. This can support maternal and newborn health, PHC service delivery, outbreak response, infection prevention, and other areas where trust, service experience, and adaptive problem-solving are central. Funding also should support dedicated learning facilitators, documentation processes, adaptive supervision, software iteration, and open-source templates for Learning Logs, Change Logs, and Empathy Tools. Current implementing partners with experience in LxD approaches can offer resources (e.g., “Learn-by-Doing Lite Toolkit”^17^ and proven tools like the Change Log, Empathy Tool, and AAR agenda templates) to facilitate such government-led efforts to strengthen existing RI and community engagement structures. Altogether, sustained facilitation capacity, leadership engagement, and alignment with routine accountability systems are essential to ensure that digital innovations can translate into operational improvements rather than parallel project structures.

Embedding LxD approaches into current interventions and processes are not without costs, and they should be regularly tracked and revisited. LxD program costing analyses, which are detailed elsewhere,^18^ indicate some additional implementation costs associated with incorporating LxD into MCH-Track (i.e., estimated financial costs of $177 USD per month and economic costs of $857 USD per month over seven months) with staff time (79%), software iterations (14%), and structured LxD activities like shadowing, documentation, and AARs (7%) accounting for the main costs. Nonetheless, cost-savings would be anticipated over time, especially as LxD investments mature and systems become more adaptive and ‘self-correcting’ in response to less effective programming.

To increase value for money, investments in ZD reduction require evidence that digital community engagement platforms include mechanisms for real-time learning, rapid adaptation, and government-owned accountability. Scaling without this adaptive layer risks expanding tools that identify missed children but fail to resolve the reasons they remain missed.

## Limitations

This program case study is subject to several limitations.

First, our work focused on implementation performance and early outcomes, and the relatively short observation period limits inference regarding changes in immunization coverage or health impact. To mitigate this, multiple implementation indicators (e.g., usability, feedback responsiveness, and workflow efficiency) were tracked over time to provide a robust assessment of early system performance; however, findings should be interpreted as indicative of implementation effectiveness rather than health impact. Measuring long-term facility-level immunization performance was outside the scope of this initial adaptation cycle, and we have noted these as important metrics for future evaluation phases.

Second, the intervention was conducted in two LGAs within Kano State, and transferability to other settings may depend on governance capacity, community structures, and digital readiness. However, the underlying LxD approach and HCD principles are inherently adaptable, and there are emerging plans to apply these methods in additional geographic settings, which will enable further evaluation of scalability and contextual generalizability.

Third, direct caregiver participation in AARs was not included in the learning process, limiting the extent to which caregiver perspectives were directly represented in the learning cycles. Future models should deliberately include caregiver representatives to strengthen community voice in learning cycles.

## Conclusion

Embedding a structured LxD approach within a digital community-engagement strategy strengthened frontline responsiveness, trust, and contextual adaptation for reaching ZD children in northern Nigeria. The findings suggest that HCD and adaptive learning systems are a critical missing component of many digital health and immunization initiatives. Institutionalizing lightweight, participatory learning mechanisms within routine immunization governance offers a scalable pathway to advance equity-focused service delivery and accelerate progress toward IA2030 to reach every child. Future research should examine long-term effects on vaccination coverage, cost-effectiveness, and national-level scale-up, while further refining adaptive learning models for diverse low-resource contexts. Furthermore, these findings suggest that LxD may offer a scalable approach for strengthening adaptive implementation across diverse health system contexts beyond just immunization. Future interventions should assess long-term effects on vaccination coverage, cost-effectiveness, and scalability across diverse settings.

## Declaration of AI Use

The authors used a generative AI tool solely for language editing to improve the clarity and readability of the manuscript. All scientific content, analyses, interpretations, and conclusions were developed and verified by the authors, who take full responsibility for the final manuscript.

## Author contributions

AAKUN: conceptualization, methodology, funding acquisition, project administration; NSN: formal analysis, investigation, methodology, validation, writing – original draft, writing – review & editing; AM: investigation, project administration; RA: data curation, investigation, project administration, visualization; NMB: writing – review & editing; MY: investigation, writing – original draft, writing - review & editing; SS: supervision, writing – review & editing; JS: methodology, supervision, writing – review & editing; PYL: supervision, writing – review & editing; KM: funding acquisition, supervision; AS: supervision, writing - review & editing; YAY: conceptualization, funding acquisition, methodology, supervision; SAI: writing - review & editing. NF: supervision, writing – review & editing. All authors reviewed and approved the final version of this article.

## Acknowledgments

We wish to sincerely acknowledge the Kano State Ministry of Health and the Kano State Primary Health Care Management Board for their leadership and support in implementing this work. We are also grateful to the development partners contributing to the immunization ecosystem in Kano State. We also appreciate our partners implementing the Zero-Dose Learning Agenda (ZDLA) in Kano State. Special appreciation goes to the community mobilizers whose dedication made field implementation possible, as well as to the Gates Foundation technical teams in Seattle and the Nigeria Country Office for their guidance and collaboration throughout the project.

## Funding

The work is funded by the Gates Foundation (INV-063621). The funder was informed of program design, data collection and analysis, decision to publish, and preparation of the manuscript, and opportunity to provide feedback. Final decisions for each - program design, data collection and analysis, decision to publish, and preparation of the manuscript - were made by program implementation team members. The corresponding author had full access to all data presented in this manuscript and had final responsibility for the decision to submit for publication.

## Competing interests

YM has received consultancy fees from the Gates Foundation from June 2025, focusing on immunization and health campaign effectiveness. SS has received funding form the Gates Foundation through McKing Consulting Corporation since 2016. JS has received consultancy fees from the Gates Foundation since 2022, largely focused on immunization, primary health care, and learn-by-doing (LxD) practices. PYL and NF are research consultants with Exemplars in Global Health at the Gates Foundation, and have received funding for work related to childhood vaccination from the Gates Foundation since 2023. KM, AS, and YAY are employees of the Gates Foundation, which funded this work. AAKUN, NSN, AM, RA, BN, and SAI declare no competing interests.

## Notes

### Competing Interest Statement

The authors have declared no competing interest.

